# Spatiotemporal changes in along-tract profilometry of cerebellar peduncles in cerebellar mutism syndrome

**DOI:** 10.1101/2021.08.03.21260011

**Authors:** Sebastian M. Toescu, Lisa Bruckert, Rashad Jabarkheel, Derek Yecies, Michael Zhang, Christopher A. Clark, Kshitij Mankad, Kristian Aquilina, Gerald A. Grant, Heidi M. Feldman, Katherine E. Travis, Kristen W. Yeom

## Abstract

Cerebellar mutism syndrome, characterised by mutism, emotional lability and cerebellar motor signs, occurs in up to 39% of children following resection of medulloblastoma, the most common malignant posterior fossa tumour of childhood. Its pathophysiology remains unclear, but prior studies have implicated damage to the superior cerebellar peduncles. In this study, the objective was to conduct high-resolution spatial profilometry of the cerebellar peduncles and identify anatomic biomarkers of cerebellar mutism syndrome.

In this retrospective case-control study, twenty-eight children with medulloblastoma (mean age 8.8 ± 3.8 years) underwent diffusion MRI at four timepoints over one year. Forty-nine healthy children (9.0 ± 4.2 years), scanned at a single timepoint, served as age- and sex-matched controls. Automated Fibre Quantification was used to segment cerebellar peduncles and compute fractional anisotropy at 30 nodes along each tract.

Thirteen patients developed cerebellar mutism syndrome. Fractional anisotropy was significantly lower in the distal segments of the superior cerebellar peduncle pre-operatively in all patients (p=0.01). Pre-operative changes in fractional anisotropy did not predict cerebellar mutism syndrome. However, post-operative reductions in fractional anisotropy were highly specific to the distal left superior cerebellar peduncle, and were most pronounced at follow-up timepoints (p=0.042,0.038), in those that developed cerebellar mutism syndrome compared to patients that did not.

High spatial resolution cerebellar profilometry identifies a site-specific alteration of the distal segment of the superior cerebellar peduncle unique to cerebellar mutism syndrome with important surgical implications in the treatment of these devastating tumours of childhood.

## Introduction

Medulloblastoma is the most common malignant brain tumour of childhood. Resective surgery plays a key therapy role but can result in post-operative cerebellar mutism syndrome (CMS) in up to 39% of cases^1^. CMS is characterized by a delayed onset of mutism, emotional lability, and cerebellar motor deficits^2^. Recovery, by way of speech dysarthria, usually occurs after several weeks, yet children are often left with longer-term linguistic and cognitive deficits^3–5^. Pathophysiological mechanisms for CMS have remained elusive, but evidence has converged on the proximal dentato-rubro-thalamo-cortical tract (DRTC)^6^. Efferent fibres of the DRTC originate in the dentate nucleus, travel through the superior cerebellar peduncle (SCP), decussate in the midbrain and ascend via the red nucleus and thalamus before projecting to widespread areas of the cerebral cortex^7–9^. Thus, damage to the SCP which contains almost all of the efferent cerebellar fibres, either from mass effect from the tumour or surgery itself, could serve as an aetiological factor^6^. The SCP may also be implicated in linguistic processes given its projections to language specific cortices such as the supplementary motor area^9,10^. At present, the role of the middle (MCP) and inferior cerebellar peduncles (ICP) in CMS is less clear, although there is some evidence of delayed structural changes in the inferior olivary nucleus following CMS^11^.

Diffusion MRI (dMRI), which interrogates voxel-wise principal diffusion directions, has offered insight into microstructure and long-range connections in the brain in CMS. The diffusion tensor is the canonical model applied to dMRI data, and yields biophysically relevant metrics such as fractional anisotropy (FA). Higher FA is seen in directionally oriented tissue microstructure, such as in white matter, due to constraints on free diffusion of water imposed by axonal structures. FA has been widely reported in many dMRI studies of children with posterior fossa tumours, and has been shown to be reduced in the SCP in children who develop CMS post-operatively compared to those who do not^12–15^. Similarly, disruptions to tractography reconstructions of the DRTC have been shown in CMS^16^. Both of these approaches, however, suffer from drawbacks.

Microstructural metrics are often averaged either across whole-brain white matter (i.e. tract-based spatial statistics^17^ as in Morris *et al*.^12^), in regions-of-interest (ROIs)^14,15^ or across the entire length of a tract^18,19^ leading to a loss of spatial information on regional variation in FA. Tractography applied in individual patients – though useful to delineate anatomy – is a qualitative tool which is difficult to truly quantify^20^. A synergistic approach is along-tract profilometry^21–23^, which enables the quantification of a chosen diffusion microstructure metric along the axis of the tract. One such toolbox, automated fibre quantification (AFQ)^22^, has yielded insights into brain-behaviour correlations for supratentorial white matter tracts. An extension has been developed to enable automated along-tract analysis of the cerebellar peduncles^24,25^, and the feasibility of its clinical application has been demonstrated^26^.

In this study, the objective was to identify biomarkers of cerebellar mutism syndrome by conducting high-resolution spatial profilometry of the cerebellar peduncles. Specifically, the study aimed firstly to validate alterations of FA at the SCPs in children who develop CMS; and secondly, to investigate the hypothesis that changes in the SCPs vary along the length of the peduncle, and across the pre- and post-operative period, thereby enhancing our currently limited understanding of the aetiology of CMS.

## Methods

### Participants

A prospectively maintained neuro-oncology database of paediatric brain tumour patients treated at Lucile Packard Children’s Hospital from 2002-2018 was retrospectively interrogated to extract clinical and demographic information on children with posterior fossa medulloblastoma (Stanford University School of Medicine / Lucile Packard Children’s Hospital Institutional Review Board #38471, 36206). Contemporaneous clinical notes on these patients were reviewed by two neurosurgeons (DY, RJ) to identify those who developed post-operative CMS. Patients were classified as having CMS if they showed the core symptom of mutism or reduced speech output in the early post-operative phase (herein referred to as CMS+). Patients were classified as not having CMS if speech output was described as normal or unchanged (CMS-). Those without explicit mention of either normal speech or speech deficit in the post-operative clinical notes were excluded from the analysis.

Imaging data were accessed for all patients, and scans were reviewed for the first post-operative year. Scans were grouped into four timepoints: pre-operative, immediate post-operative (< 7 days of tumour resection); early follow-up (1-4 months); and late follow-up (> 9 months). Patients with dMRI acquisition for at least one timepoint were included in the study. Scans acquired between 4 and 9 months post-operatively were not included so as to increase the distinction between the late follow-up timepoints. Twenty patients had more than one scan within a single timepoint. In this situation, the scan which was technically superior (i.e. no movement artefact or missing sequences) was selected. When both scans were comparable in quality, the scan closest to the operation date for time points 1-3 or the scan closest to the 12-month post-op date for time point 4 was chosen. Supplementary Figure 1 shows dMRI availability in the entire cohort of eligible patients.

Timepoint-specific age- and sex-matched healthy controls were selected from a cohort of 113 children and adolescents who presented at Lucile Packard Children’s Hospital from 2010-2017. In all cases, these children had non-specific indications for scanning, such as family history of aneurysms or cancer risks, headaches, dizziness, nausea, but had normal structural MRI scans, as reviewed by a paediatric neuroradiologist (KWY), and no known systemic conditions affecting the brain, or psychiatric or developmental disorders, based on medical record review by a paediatrician (HMF). Each control participant had a single scan for analysis. Participant selection and results of along-tract cerebellar profilometry of this cohort of healthy children are detailed elsewhere^25^. If two control candidates had the same age, both were included in the comparison group in order to enrich the control dataset; if a control candidate had an identical age to a patient, but different sex, the subject was included; sex-matching was performed at a group level, ensuring balance in sexes. Table 1 shows the number of subjects included at each timepoint.

**Table 1.** Demographics of included study participants.

| | | Scan timing (months post-op) | n | Age (y, mean±S.D.)* | Male % | CMS+ | CMS present % |
| --- | --- | --- | --- | --- | --- | --- | --- |
| Pre-op | Patients | 0 | 13 | 7.68 ± 3.31 | 69.2 | 6** | 46.2 |
| | Controls | N/A | 25 | 7.79 ± 4.04 | 68.0 | N/A | N/A |
| Post-op | Patients | 0.08 ± 0.04 | 14 | 9.09 ± 4.13 | 78.6 | 7 | 50.0 |
| | Controls | N/A | 27 | 8.48 ± 4.62 | 66.7 | N/A | N/A |
| Early FU | Patients | 2.93 ± 0.81 | 13 | 9.78 ± 3.94 | 76.9 | 6 | 46.2 |
| | Controls | N/A | 20 | 8.97 ± 4.03 | 65.0 | N/A | N/A |
| Late FU | Patients | 12.5 ± 2.92 | 18 | 10.3 ± 4.00 | 66.7 | 11 | 61.1 |
| | Controls | N/A | 26 | 10.7 ± 3.76 | 61.5 | N/A | N/A |

49 unique control subjects were included overall; 28 unique patients were included overall.
\*, no significant differences in age between patients and controls at each timepoint (all t test $p > 0.572$ ); \*\*, diagnosis of CMS applied post-operatively; CMS+, cerebellar mutism syndrome present; FU, follow up.

### MRI acquisition

MRI scans were acquired on a 3T GE MR750 Discovery (GE Healthcare, Waukesha, WI, USA) using an 8-channel head coil. Children aged up to 6 years old were sedated under general anaesthesia; some children aged 6–8 years were sedated based on individual maturity level and ability to tolerate the MRI exam. Both high-resolution T1-weighted (3D SPGR, TR = 7.76 ms, TE = 3.47 ms, FOV = 240 × 240 mm^2^, acquisition matrix = 512 × 512, voxel size = 0.4688 × 0.4688 × 1 mm^3^, orientation = axial) and diffusion-weighted images were acquired as part of the paediatric brain MRI protocol. Diffusion data were collected with a twice-refocused GRAPPA DT-EPI sequence (TR = 4000–6000 ms depending on slice coverage, TE = 76.59 ms, FOV = 240 × 240 mm^2^, acquisition matrix = 256 × 256, voxel size = 0.9375 × 0.9375 × 3 mm^3^) using a b-value of 1000 s/mm^2^ sampling along 25 isotropically distributed diffusion directions. One additional volume was acquired at b = 0 at the beginning of each scan.

### Image preprocessing

After file conversion from Dicom to NIFTI using *dcm2niix*^27^, dMRI data were preprocessed using the open-source software mrDiffusion (github.com/vistalab/vistasoft/mrdiffusion) implemented in Matlab R2017b (Mathworks, Natick, MA, USA). The b0 image was registered to the patient’s anatomical T1-weighted image, which had been centred on the anterior commissure and aligned to the anterior-posterior commissural plane. The combined transform that resulted from motion correction and alignment to the T1-weighted image was applied to the raw data (as well as the diffusion gradient tables), and the transformed images were resampled to 2 × 2 × 2mm isotropic voxels. The diffusion tensor was fitted using a ‘least squares’ method, and colour FA maps co-registered to the T1-weighted image were visually checked to confirm alignment.

Relative head motion was quantified by calculating the magnitude of motion correction (in voxels) applied in the pre-processing step, for each dMRI volume relative to the previous volume. This was done at each timepoint for each subject. For each timepoint, the mean number of subject volumes with more than 1 voxel of relative motion was calculated. If any subject had more than this group mean+3SD number of volumes with >1 voxel of relative motion, that subject was discarded (1 subject at the late follow-up timepoint).

Global FA was calculated for all subjects included at each timepoint. Subjects with a global mean FA of more than 3 SD from the group mean were excluded from the analysis (2 subjects at the late follow-up timepoint only).

### White matter tract identification

The open-source toolbox Automated Fibre Quantification (AFQ)^22^ implemented in Matlab R2017b was used to perform tractography for each subject in an automated fashion. AFQ first performs whole-brain tractography, then segments fibre tracts based on atlas ROIs warped into the subject’s native space. These fibre tracts are then refined and clipped to the ROIs before quantification of diffusion metrics at a user-defined number of locations, or ‘nodes’, along the tract. A detailed description of cerebellar AFQ methodology is provided by Bruckert *et al*.^25^. Briefly, deterministic whole-brain tractography using a streamlines tracking technique^28–30^ was seeded from a white matter mask defined as voxels with an FA value greater than 0.15; tracking was terminated in voxels with an FA below 0.1^25^. Fibres shorter than 20mm and longer than 250mm were discarded from the whole-brain tractogram. Non-linear registrations were used to warp ROIs from MNI space into subject space. The inferior cerebellar peduncles (ICP), superior cerebellar peduncles (SCP) and middle cerebellar peduncle (MCP) were segmented if fibres from the whole-brain tractogram passed through the relevant ROIs. The tract is then clipped between the seed and waypoint ROI, and divided into 30 nodes. The robust mean position of the tract was computed by estimating the three-dimensional Gaussian covariance of the sample points and removing fibres that were either more than 4 standard deviations away from the mean position of the tract or that differed more than 1 standard deviation in length from the mean length of the tract for relatively short cerebellar pathways. Fibre tracts were then clipped to begin and end at the ROIs from which they were created.

Fibre renderings of each subject’s tracts, at each timepoint, were visually inspected; those which did not conform to known anatomical configurations of cerebellar peduncles were discarded (Table 2). None of the controls’ fibre groups were discarded as these had previously been through a rigorous quality control process^25^. For each tract in each subject, FA was summarized at 30 equidistant nodes by taking a weighted average of all streamlines at that node.

**Table 2.**
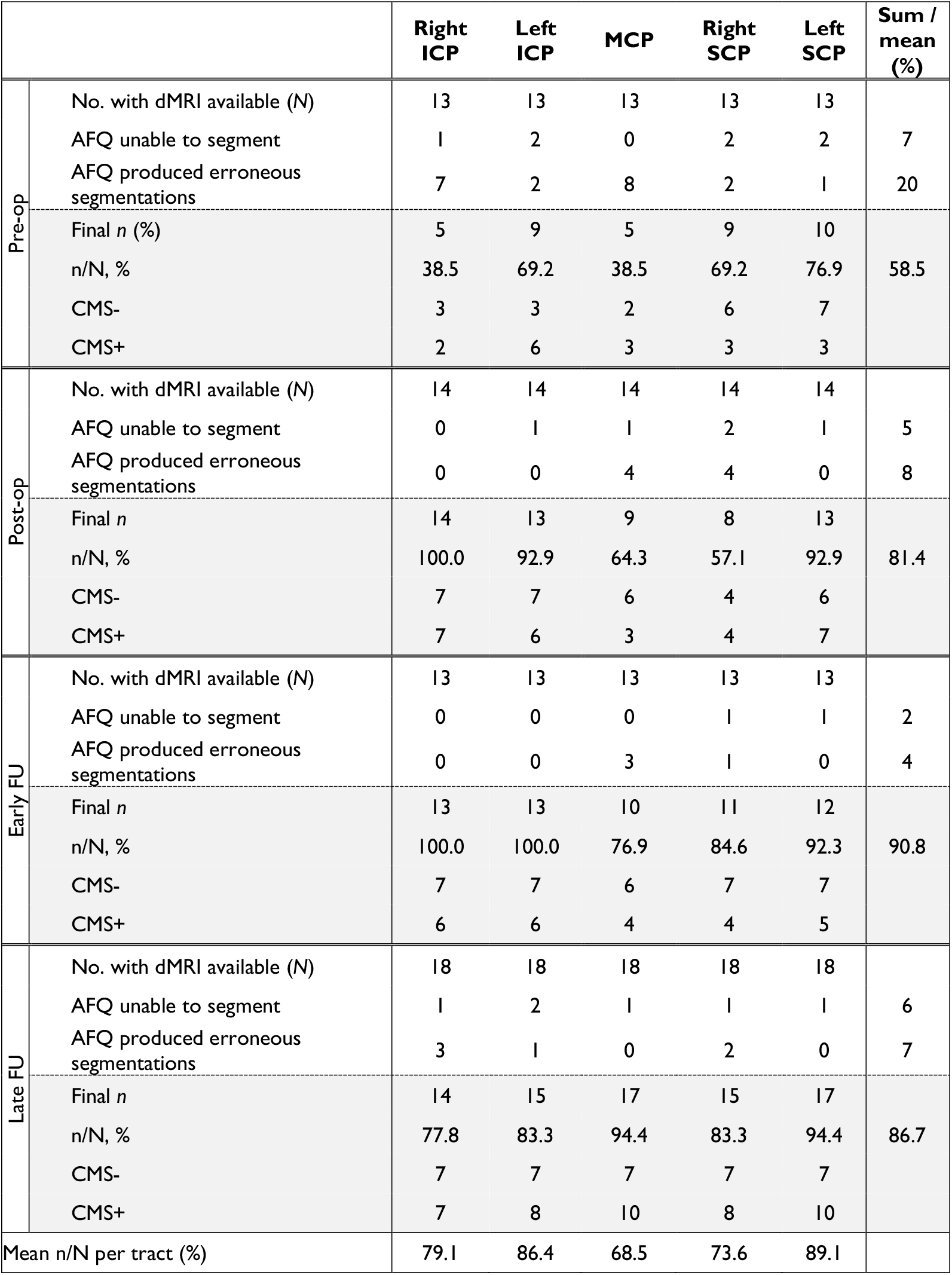
Breakdown of fibre groups included by timepoint in patients with medulloblastoma.

### Statistical Analysis

Tractography results were imported into R (v3.6.1; R Core Team, 2017) for further analysis and graphics were created using the ggplot2^31^, rstatix^32^ and ggpubr^33^ packages. A pre-specified α of 0.05 was chosen. Results at each timepoint and tract underwent independent statistical analysis. In order to test for group differences in along-tract profilometry between controls, CMS+ and CMS-, one-way ANOVA tests were computed on a node-by-node basis for each cerebellar peduncle and timepoint, generating a p-value at each node (p_var_). A nonparametric permutation-based method^34^ was used to control for the 30 comparisons along the tract. This procedure produced a family-wise error corrected cluster size and a critical p-value (p_min_) for each of the candidate tracts. Differences in nodal FA values were considered significant if either of two criteria were satisfied: 1) p_var_ < α at a sufficient number of adjacent nodes to meet the criteria for a family-wise error corrected cluster size (i.e. if cluster size = 8; ≥8 adjacent nodes required p_var_ < α); or, 2) for clusters not meeting the required cluster size or for single nodes, p_var_ < p_min_.

An important consideration in this work was to test for group differences in FA between CMS+ and CMS-groups. In order to achieve this step, each patient’s FA values were standardised to Z-scores based on the age- and sex-matched controls’ summary scores at each node, for corresponding timepoints and tracts. Each tract was then segmented into thirds in order to make subsequent statistical testing more concise. The proximal third in the SCPs corresponded to nodes closest to the dentate nucleus, and the distal third corresponded to nodes closest to the midbrain. The ICP nodes closest to the inferior olivary nucleus corresponded to the proximal third, and those in the cerebellum were the distal third. MCP nodes ascended from right to left along the extent of the tract. For each cerebellar peduncle and timepoint, patients’ Z-scores were compared using a two-way mixed ANOVA, with a between-subjects factor of group (CMS+ vs CMS-), and a within-subjects factor of tract segment (1^st^ third, 2^nd^ third, 3^rd^ third). *Post-hoc* false-discovery rate adjusted^35^ pairwise t-tests were used to compare significant interactions between CMS status and tract segments.

### Data availability

The AFQ toolbox is freely available for use within Matlab and Python environments (https://github.com/yeatmanlab/AFQ). Anonymised data reported within this paper are available upon reasonable request to the senior author.

## Results

A total of 28 patients with medulloblastoma were included in this study (13 CMS+, 15 CMS-), along with a total of 49 age- and sex-matched healthy controls. Twenty-one patients (70%) had dMRI scans available at multiple timepoints. Twenty-seven patients (90.0%) had midline tumours of the vermis or fourth ventricle; three patients had tumours in the cerebellar hemispheres. Patient inclusion in different timepoints can be seen in Supplementary Figure 1. Table 1 shows demographic information of the included patients and their age- and sex-matched controls at each timepoint. The proportion of children with CMS at each timepoint varied from 46.2-61.1%. At one year follow-up, symptoms of CMS had resolved in 9/13 (69.2%). The remaining four children had ongoing speech-language pathology treatment.

### Fibre tracking

Automated fibre segmentation successfully identified the cerebellar peduncles in the majority of participants; an example patient is shown in Figure 1. Some participants’ fibres were excluded after visual inspection. While different fibres were excluded at different timepoints, a larger proportion was discarded at peri-operative timepoints owing to distorted brain anatomy due to the tumour mass effect or resection cavity. Table 2 provides a detailed breakdown of fibres included for patients at each timepoint.

**Figure 1.**
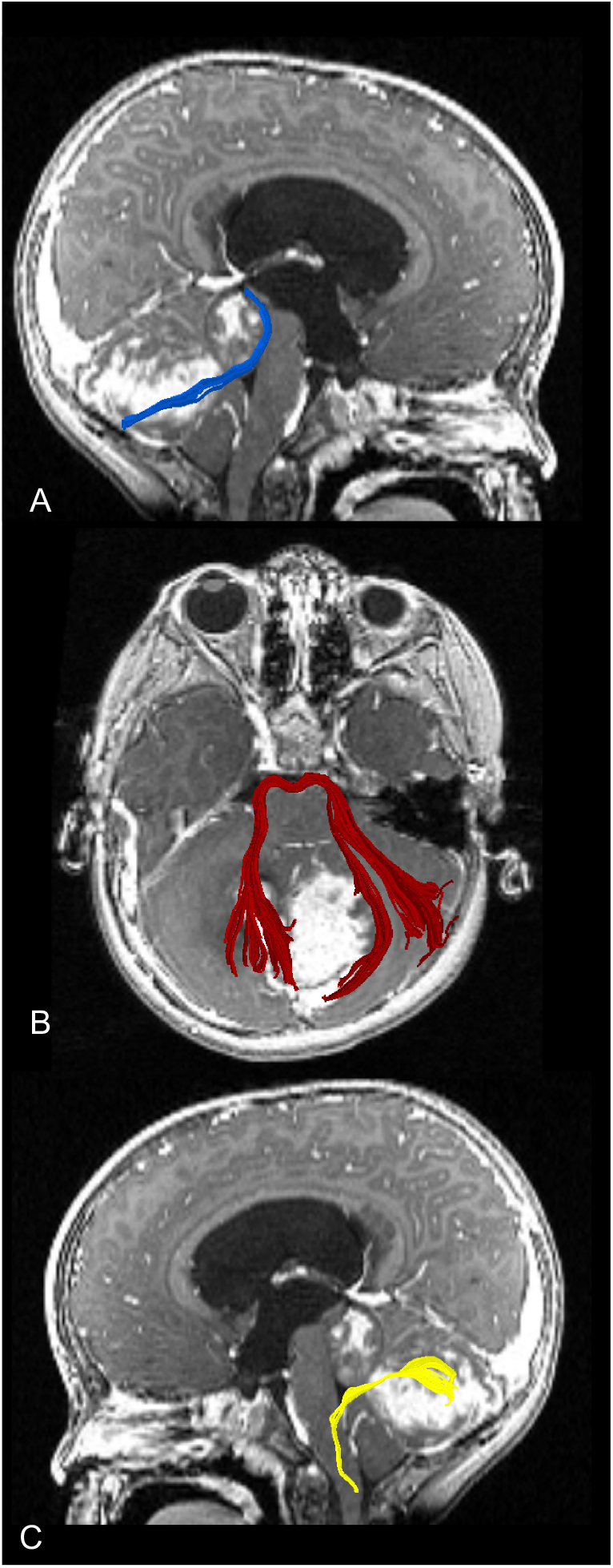
Example of AFQ-segmented cerebellar peduncles shown on T1-weighted post-contrast images of a representative subject with medulloblastoma. A, parasagittal section showing the right superior cerebellar peduncle in blue; B, axial section showing middle cerebellar peduncle in red; C, parasagittal section showing the left inferior cerebellar peduncle in yellow.

### Along-tract profiles of cerebellar peduncles

The results of both analysis approaches (along-tract profiles with permutation-based multiple comparison corrected significance tests; and mixed ANOVAs, *post-hoc* tests, and z-score plots) are described in tandem with reference to each tract, below. Mixed ANOVA results are reported in full in Supplementary Table 1.

### Superior cerebellar peduncles

#### Left SCP

The left SCP showed the greatest effect sizes in group comparisons of along tract FA profiles. Pre-operatively, the difference in distal left SCP FA between controls and patients was statistically significant (Figure 2A/i), but patients could not be distinguished based on their CMS status (a classification which was applied retrospectively, following tumour resection). Immediately following surgery, along-tract profiles showed that left SCP FA increased in CMS-, but remained lower, especially distally, in CMS+ (Figure 2A/ii). At early follow-up scanning, there were significant differences in left SCP FA between controls, CMS+ and CMS-(Figure 2A/iii). At the late follow-up timepoint, there were statistically significant group differences at almost all nodes along the tract (Figure 2A/iv).

**Figure 2.**
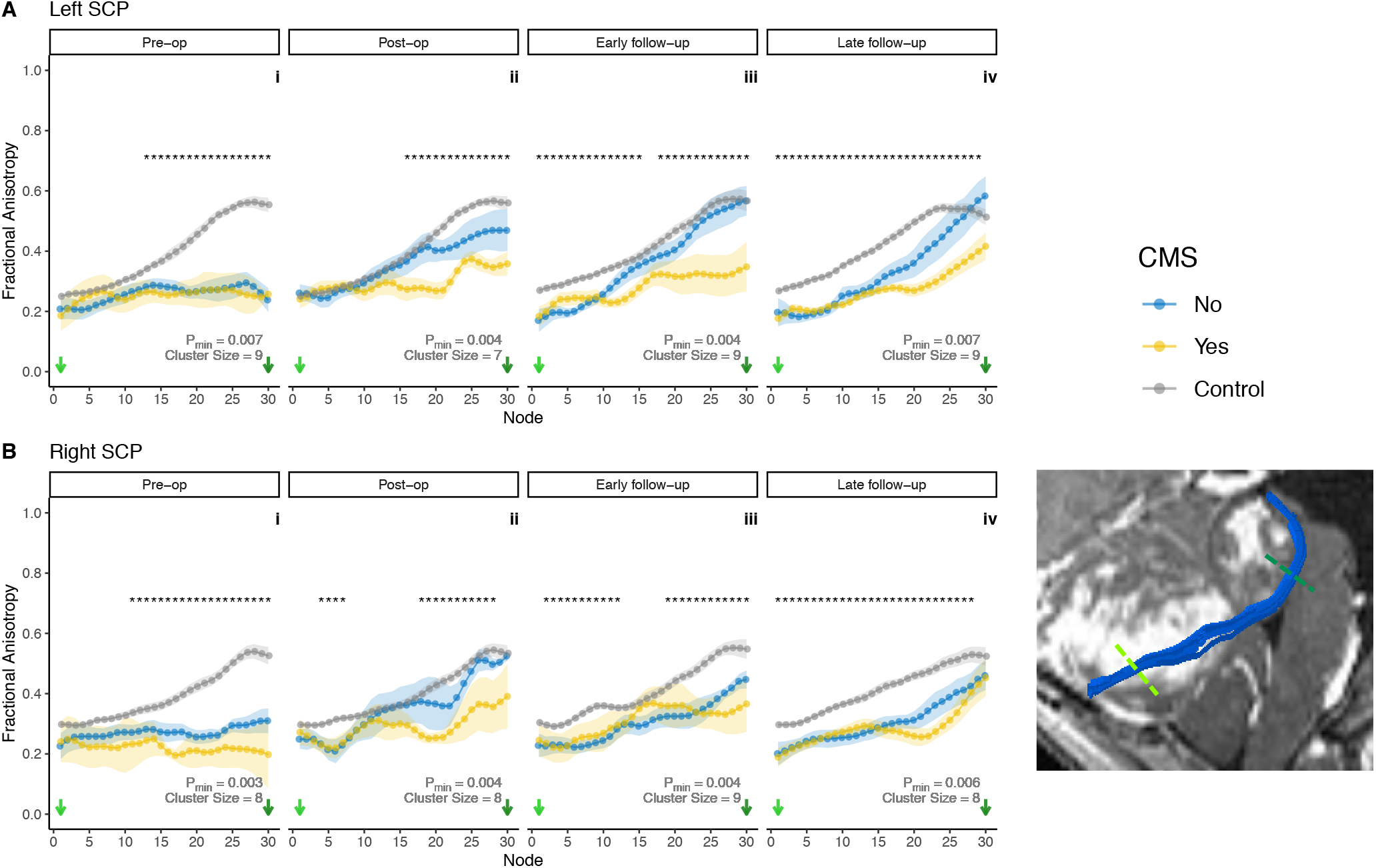
Along-tract profilometry results for the superior cerebellar peduncles (SCP) in patients and controls across four timepoints, pre-op (i) to late follow-up (iv). **A**, left SCP; **B**, right SCP. Points indicate group mean at each node; ribbon indicates standard error of the mean per group. * indicate significant group differences as described in the ‘Statistical Analysis’ section. Cluster size and p_min_indicated for each tract and timepoint. Bottom right inset shows a representative example of a segmented SCP (blue) in a patient with medulloblastoma. Coloured dashed lines on inset correspond to node number on plots.

At the pre-operative timepoint (Figure 3A/i), mixed ANOVA analysis in tract segments of the left SCP showed no significant main effect of CMS status alone (F(1,8)=0.003, p=0.957). The interaction between tract segment and CMS status was not statistically significant (F(2,16)=0.229, p=0.798). There was a significant main effect of tract segment on z-score (F(2,16)=7.361, p=0.005), with significant differences also seen on *post-hoc* testing between distal vs proximal two thirds of the left SCP (p=0.014 for both).

**Figure 3.**
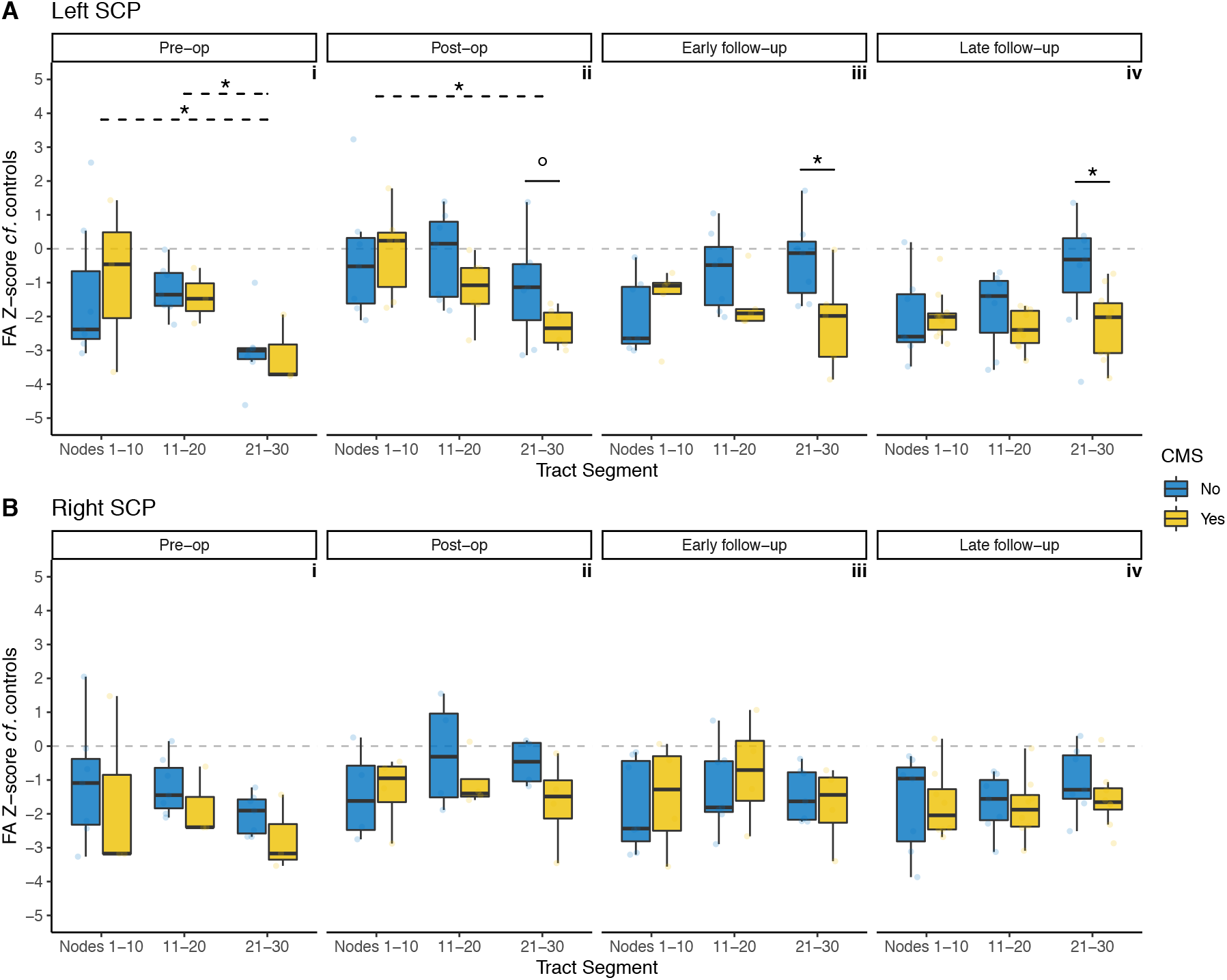
Z-score of FA for patients with CMS (yellow) and without (blue), compared to controls, within tract segments (proximal / middle / distal thirds) of the superior cerebellar peduncles (SCP). **A**, left SCP; **B**, right SCP. Grey dotted line indicates z = 0. False discovery rate adjusted p values: °, p<0.1; *, p<0.05. Solid lines indicate *post-hoc* pairwise significance tests between CMS groups within tract segments; dotted lines indicate *post-hoc* pairwise significance tests between tract segments.

At the post-operative timepoint (Figure 3A/ii), mixed ANOVA analysis in the left SCP showed a persistent main effect of tract segment (F(2,22)=11.5, p<0.001), underpinned by a significant difference between proximal and distal SCP (p=0.026). There were no significant effects of CMS (F(1,11)=0.992, p=0.341) or its interaction with tract segment (F(2,22)=2.54, p=0.102). *Post-hoc* testing showed that while distal left SCP FA was lower in the CMS+ group, this did not reach a level of statistical significance (p=0.1).

At early follow-up scanning (Figure 3A/iii), mixed ANOVA analysis in the left SCP showed a significant interaction between CMS status and tract segment (F(2,20)=6.72, p=0.006). *Post-hoc* testing revealed that the interaction was due to statistically significant group differences at the distal SCP (p=0.042). There were no significant main effects of CMS alone (F(1,10)=1.80, p=0.209) or tract segment (F(2,20) = 1.99, p=0.163) at this timepoint.

At late follow-up scanning (Figure 3A/iv), mixed ANOVA analysis in the left SCP showed a persistent significant interaction between tract segment and CMS (F(2,30)=6.07, p=0.006), which was again was driven by a statistically significant group difference at the distal left SCP (p=0.038). There was a significant main effect of tract segment (F(2,30)=4.03, p=0.028), but no significant pairwise differences on *post-hoc* testing. There was no significant effect of CMS alone (F(1,15)=2.16, p=0.162).

#### Right SCP

Along-tract FA profiles of the right SCP showed similar patterns to its contralateral counterpart. Pre-operatively, there was a statistically significant difference between groups towards distal nodes of the right SCP (Figure 2B/i). Post-operatively, there were significant group difference clusters at the proximal and distal right SCP (Figure 2B/ii). At early follow-up scanning, these clusters were larger (Figure 2B/iii). At late follow-up scanning, there were statistically significant group differences at almost all nodes along the tract (Figure 2B/iv).

Mixed ANOVA analysis along the right SCP did not demonstrate any statistically significant interactions of CMS and tract segment at any timepoint (Figure 3B). In particular, the lower FA seen pre-operatively in the distal R SCP (Figure 2B/i) did not reach statistical significance in the mixed ANOVA analysis (Figure 3B/i). There were also no significant effects of CMS status or tract segment alone, at any timepoint, in the right SCP.

### Inferior Cerebellar Peduncles

#### Left ICP

Small clusters of significant group differences in along-tract FA profiles were seen the left ICP at pre-operative, post-operative and late follow-up timepoints (Figure 4A).

**Figure 4.**
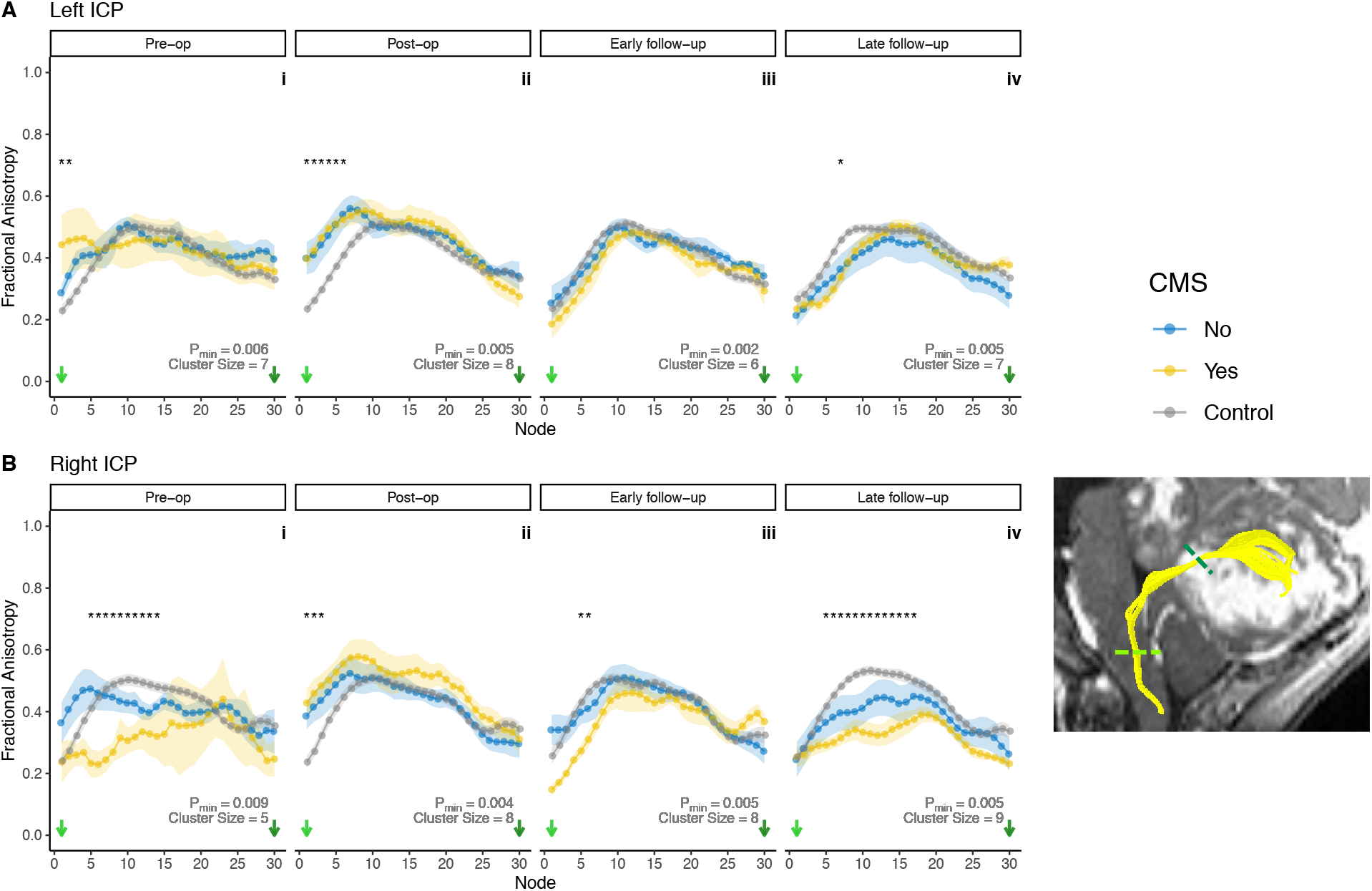
Along-tract profilometry results for the inferior cerebellar peduncles (ICP) in patients and controls across four timepoints, pre-op (i) to late follow-up (iv). **A**, left ICP; **B**, right ICP. Points indicate group mean at each node; ribbon indicates standard error of the mean per group. * indicate significant group differences as described in the ‘Statistical Analysis’ section. Cluster size and p_min_indicated for each tract and timepoint. Bottom right inset shows a representative example of a segmented ICP (yellow) in a patient with medulloblastoma. Coloured dashed lines on inset correspond to node number on plots.

Mixed ANOVA analysis along the left ICP (Figure 5A) did not reveal any statistically significant effects of CMS or its interaction with tract segment, at any timepoint. This analysis did reveal statistically significant main effects of tract segment in determining FA z-score for the left ICP at all timepoints apart from pre-operatively (post-operative F(2,22)=11.8, p<0.001); early follow-up F(2,22)=3.78, p=0.039; late follow-up F(2,26)=3.91, p=0.033). *Post-hoc* pairwise comparison tests reached statistical significance at the post-operative timepoint between the first and second (p=0.003), and first and third (p=0.002) segments of the tract (Figure 5A/ii).

**Figure 5.**
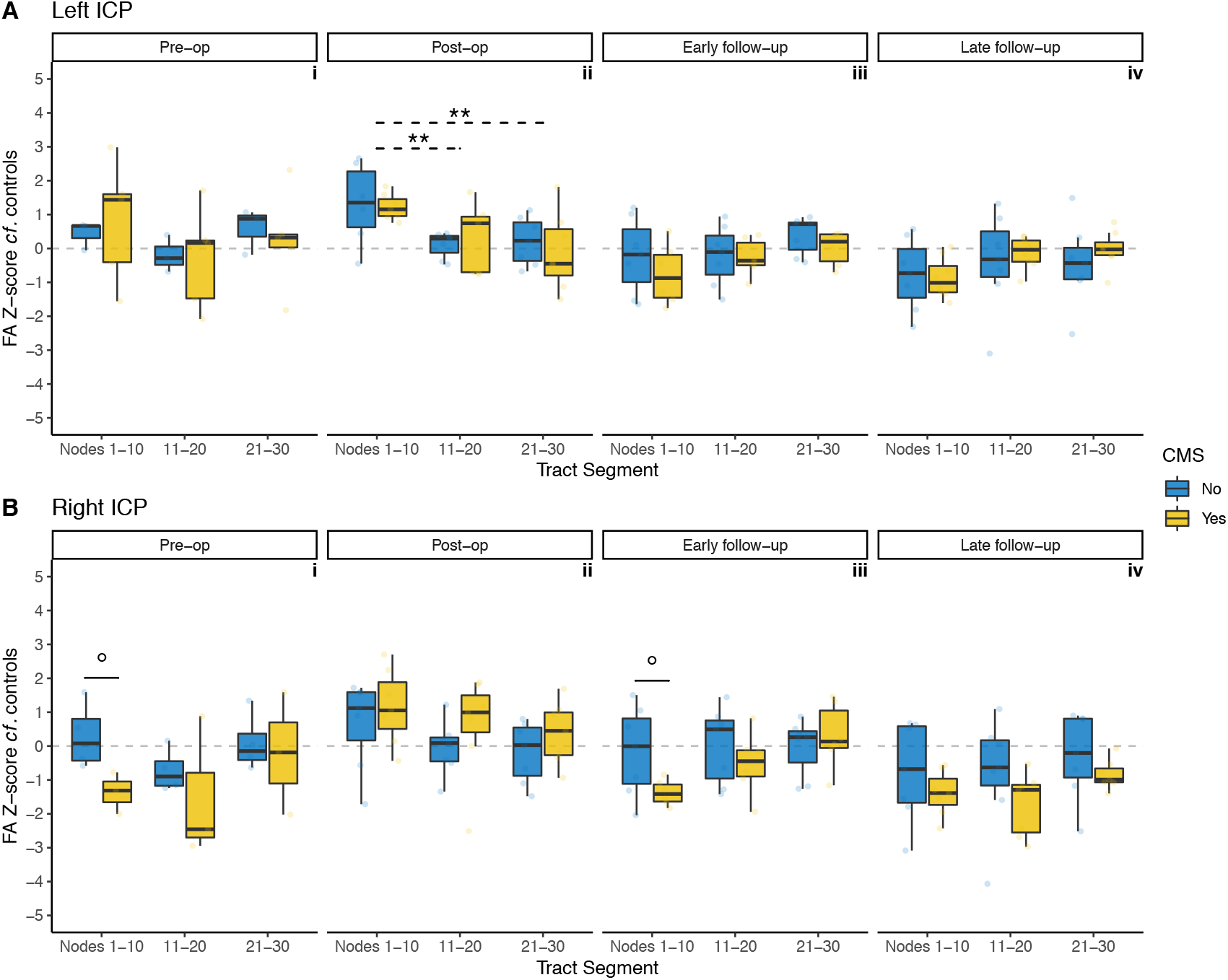
Z-score of FA for patients with CMS (yellow) and without (blue), compared to controls, within tract segments (proximal / middle / distal thirds) of the inferior cerebellar peduncles (ICP). **A**, left ICP; **B**, right ICP. Grey dotted line indicates z = 0. False discovery rate adjusted p values: °, p<0.1; *, p<0.05. Solid lines indicate *post-hoc* pairwise significance tests between CMS groups within tract segments; dotted lines indicate *post-hoc* pairwise significance tests between tract segments.

#### Right ICP

Pre-operative along-tract FA profiles of the right ICP showed a significant cluster of group differences in the proximal tract (Figure 4B/i). Post-operatively and at early follow-up scanning, smaller clusters of group differences in along-tract FA results were apparent (Figures 4B/ii and iii). At late follow-up scanning, the same cluster observed at the pre-operative timepoint was again apparent (Figure 4B/iv)..

Mixed ANOVA analysis in the right ICP at the pre-operative timepoint (Figure 5B/i) did not reveal statistically significant main effects of tract segment (F(2,10)=3.81, p=0.059) or CMS alone (F(1,5)=1.25, p=0.314). There was also no significant main effect for interaction of tract segment with CMS (F(2,10)=1.58, p=0.254). Although *post-hoc* pairwise comparison testing at the proximal right ICP showed markedly lower z-scores in CMS+ than CMS-, this did not reach statistical significance (p=0.053).

Post-operatively, mixed ANOVA analysis in the right ICP (Figure 5B/ii) showed a significant main effect of tract segment (F(2,24)=7.52, p=0.003), although pairwise comparisons of tract segment at this timepoint did not survive multiple comparison correction. There was no significant main effect of CMS status (F(1,12)=1.12, p=0.310) or its interaction with tract segment (F(2,24)=0.068, p=0.934).

At early follow-up, mixed ANOVA analysis in the right ICP (Figure 5B/iii) showed a significant main effect for tract segment (F(2,22)=6.56, p=0.006) and its interaction with CMS (F(2,22)=4.79, p=0.019). *Post-hoc* pairwise testing at the proximal right ICP showed lower z-scores in CMS+ than CMS-, but this did not reach a level of statistical significance (p=0.051). CMS alone had a non-significant main effect at this timepoint (F(1,11)=1.03, p=0.332).

At late follow-up scanning, mixed ANOVA analysis along the right ICP (Figure 5B/iv) did not show significant main effects of CMS (F(1,12)=1.82, p=0.202), tract segment (F(2,24)=3.30, p=0.054), or their interactions (F(2,24)=0.263, p=0.771).

### Middle Cerebellar Peduncles

The MCP tract profiles did not show any statistically significant differences between groups at any individual node, and therefore no significant clusters of group differences. There were no significant main effects of CMS, tract segment or their interactions at any timepoint. Results for the MCP are shown in Figure 6.

**Figure 6.**
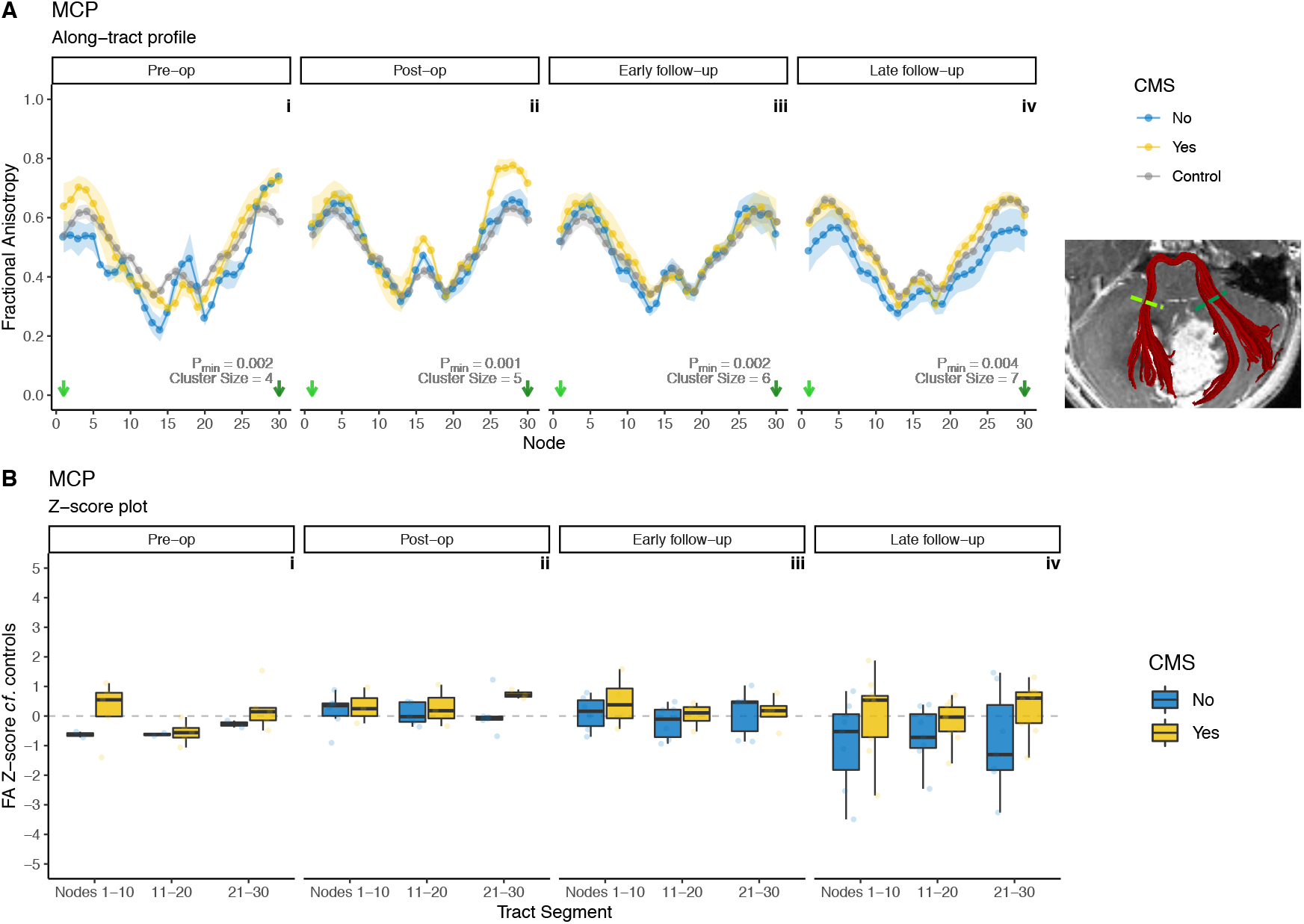
**A**, Along-tract profiles of the middle cerebellar peduncle (MCP) in patients and controls, across four timepoints, pre-operative (i) to late follow-up (iv). There were no statistically significant differences between groups at any individual node. Inset shows a representative example of a segmented MCP (red) in a patient with medulloblastoma. Coloured dashed lines on inset corresponds to node number on plots. **B**, Z-score of FA for patients with CMS (yellow) and without (blue), compared to controls, within tract segments (proximal / middle / distal thirds) of the middle cerebellar peduncles (MCP). Grey dotted line indicates z = 0. There were no statistically significant differences with respect to CMS status or tract segment.

## Discussion

In this retrospective cohort study of children undergoing resective surgery of posterior fossa medulloblastoma, automated along-tract profilometry was used to investigate spatial changes in FA in the cerebellar peduncles in a cross-sectional fashion at four timepoints over the first post-operative year. Along-tract profilometry, an advanced processing method which is highly sensitive to local changes in underlying dMRI metrics, demonstrated visually appreciable differences in FA between controls, CMS+, and CMS-. Analysis of standardised FA values, used to compare CMS+ to CMS-, found reductions in post-operative FA in CMS+ which were highly specific to the SCPs; these group differences were observed up to one year after surgery and were particularly localised to the distal left SCP.

### dMRI metrics in CMS

The first report of altered DTI metrics in children with CMS^12^ studied a group of 26 patients with medulloblastoma, half of whom developed CMS post-operatively. Tract-based spatial statistics (TBSS)^17^ was used in order to assess group differences in FA. This approach showed significantly reduced FA in bilateral SCPs, as well as in supratentorial white matter subtending the right angular gyrus, the left superior frontal gyrus, and also in the columns of the fornix^12^. Subsequently, investigators examined other dMRI metrics in CMS. Law *et al*., 2012 showed significant group differences in mean, axial, and radial diffusivity, but not FA, in right cerebellar white matter based on deterministic tractography in a group of 51 children with heterogeneous posterior fossa tumours (17 of whom developed CMS), and 28 healthy controls^19^. Qualitative assessment of direction-encoded colour FA maps^36^ and intra-operative diffusion-weighted MRI^37^ suggested SCP damage in CMS. Tract volumes of fronto-cerebellar fibre tracts have also been shown to be diminished in children with CMS^18^.

A distinctive feature of the present study is the investigation of cerebellar white matter tracts in cerebellar mutism at multiple timepoints, from pre-surgical to 1-year post-operative. The aforementioned studies all concern the application of dMRI at a single, post-operative timepoint. Given the striking temporal course of CMS semiology – beginning days after tumour resection and usually receding after a few months – longitudinal MRI studies could offer important insight in this clinical population. In a cohort of 14 posterior fossa tumour patients unstratified by CMS diagnosis, significant decreases in SCP FA were seen at early follow-up imaging, with a later increase accompanied by dentate nucleus volume loss^13^. A longitudinal study at three timepoints (pre-op, post-op and 1 year follow-up) showed statistically significant post-operative reductions in FA of both SCPs which persisted up to 1 year^14^. A similarly designed study showed post-operative reductions in FA in the left SCP only, and these changes were persistent in patients with ongoing ataxia at delayed follow-up imaging, but not in those with ongoing symptoms of mutism^15^. Neither of the latter two studies demonstrated pre-operative changes in FA at the SCP in patients who went on to develop CMS^14 15^,.

The present study identifies no statistically significant differences in pre-operative FA of any cerebellar peduncle with respect to subsequent CMS status. The pre-operative divergence in right SCP FA between CMS+ and CMS-at the pre-operative timepoint (Figure 2B/i) is intriguing though it did not achieve statistical significance. Cumulative evidence thus far indicates that FA in an insufficiently sensitive pre-operative biomarker of CMS risk. Similar findings have been found for T2-weighted signal change^38^. However, it is possible that within the sample is a subset of patients for whom FA of the SCPs might be a useful biomarker. Therefore, further study with larger sample sizes is warranted.

### Spatial sensitivity of along-tract profilometry in CMS

The present study used along-tract profilometry, an offshoot of dMRI post-processing which enables the sampling of dMRI metrics along the long axis of a tract, rather than within regions of interest on a voxel-wise basis. Several implementations of this processing method exist^21,23^, and here we report a novel application of one of the earliest such toolboxes, AFQ^22^. Initially devised as a means of segmenting major supratentorial white matter tracts, a later extension to the toolbox added cerebellar peduncle segmentation^24,25^, and this application has been used to study reading ability in healthy children^39^, locomotor adaptation in healthy adults^40^ and cerebellar microstructure in preterm-born adolescents^26^.

The AFQ pipeline offers major advantages for segmenting white matter pathways in a peri-operative neurosurgical population. The method aligns the patient’s scan with a template only for the purpose of automatically placing ROIs in the patient’s native scan. After that step, however, tractography takes place in native space. Despite potential challenges to segmentation and tracking at the peri-operative timepoints due to mass effect distortions, the AFQ approach generally performed well (Table 2). Given the plethora of increasingly complex dMRI processing tools available, efforts are underway to standardise dMRI pipelines^41^, and AFQ dovetails well with this mission. Ultimately, an automated pipeline avoids manual and subjective ROI placement and is likely more robust to interobserver bias.

A further advantage of the present methodology is that it represents a significant advance in describing the location-dependent changes in FA at the SCP FA. Evidence from several studies^12,14–16^ has converged on the SCP as being implicated in CMS. However, these studies do not spatially localize changes within the SCP. Inspection of Figure 2 of Morris *et al*.’s work^12^ shows that the FA changes were seen across the entirety of both SCPs, with some retrograde extension into the cerebellar white matter. SCP ROIs in Vedantam *et al*.^15^ were placed on three axial slices, with FA results averaged across these slices. McEvoy *et al*.^14^ used ROIs from a co-registered DTI atlas^42^, with FA averaging within the ROI. The present study identifies site-specific alteration of FA in the distal SCP as it approaches the midbrain rather than its more proximal segment as it exits the cerebellar parenchyma. Furthermore, assessing these changes over four timepoints (including pre-operatively), with at least 13 patients at each being compared to a robust number of age-and sex-matched controls, makes the present study a richly sampled dataset in comparison to other series.

Complementary analysis techniques were used to assess differences in along-tract FA. First, differences in mean FA of each group (controls, CMS+ and CMS-) were compared at each of thirty nodes along the segmented tracts, using one-way ANOVA. Due to the high degree of auto-correlation between adjacent nodes, a non-parametric permutation based method was used to generate a p value (p_min_) adjusted for multiple comparisons, as well as a minimum cluster size for truly significant group differences. Figures 2 and 4 show that significant clusters were concentrated mainly in the SCPs, with smaller clusters in the ICPs. There were no significant group differences in along-tract FA for the MCP.

The present study also converted the patients’ FA values to z-scores based on the controls’ summary statistics of FA at each node. This method allowed a comparison of the magnitude of difference in FA between the patient cohort and the control children; and, crucially, an assessment of differences within the patient cohort with respect to CMS status. The z-score of distal SCP FA in CMS+ was -2 (2 standard deviations lower than the control mean); whereas CMS-had z-scores in this location close to 0 (similar to mean FA of controls at those nodes). Crucially, mixed ANOVA showed a significant main effect of interaction between CMS and tract segment at these two timepoints (p=0.006 for both), driven by statistically significant differences at the distal SCP.

The present study suggests that in CMS, changes in white matter are highly localized at the rostral portion of the SCP – in other words, the part of the SCP which forms the lateral walls of the fourth ventricle. If damage to the SCP in CMS is putatively caused at the time of surgery, neurosurgeons performing tumour resections in this location will wisely treat this area with extreme caution, particularly as fibres to the supplementary motor area, with its key involvement in speech generation, are thought to reside in the medial edge of the SCP^9^ where they are at risk from surgical manipulation. Although mixed ANOVA found these spatial differences to be significant at early and late follow-up timepoints only, along-tract analyses also indicated evidence of group differences in the distal SCP at the post-operative timepoint. These findings suggest that surgical damage contributes to acute, as well as chronic, changes in FA in the distal SCP.

Reduced FA in the left SCP in CMS+ compared to controls and CMS-patients may result from greater directional incoherence – or, in a biophysical sense, less uniformly arrayed axons. However, FA encodes both microscopic diffusion processes (i.e. related to the density of fibres) as well as orientational information. More advanced models of the diffusion signal, such as the spherical mean technique^43^, are able to distinguish between these two components and can contribute precise estimates of changes in orientational tissue properties.

Combining spherical mean technique modelling with AFQ profilometry could yield further insights into the fine-grained microstructural properties of cerebellar peduncles in children with CMS.

### Limitations

This study effectively builds on previous work and represents a richly-sampled analysis of CMS patients using DTI. A limitation is the modest sample size for some groups and timepoints (e.g. pre-operatively, the right ICP and MCP analysis only contained 5 patients). The numbers of viable tracts included in the analyses were unavoidably reduced at the stage of visual inspection of AFQ fibre segmentations. This problem was most commonly seen at the peri-operative timepoints, appreciably due to anatomical distortion from tumours in pre-operative, and resection cavities in post-operative scans. By the early follow-up timepoint, however, 90.8 of fibre groups segmented by AFQ were found to be viable. Across all timepoints, AFQ segmentation of the left SCP was the most robust, deemed anatomically accurate in 89.1% of cases.

The lack of an objective determination of CMS status is a perennial issue in studies reporting neuroimaging correlates of the syndrome and this criticism applies to this study. The diagnosis of CMS were applied based on retrospective review of case notes. Every attempt was made to limit potential bias by double review of notes by trained neurosurgeons, and in cases where no clear contemporaneous description of mutism was annotated, these were classified as having indeterminate CMS status and excluded from the imaging analyses. Moreover, careful testing longitudinally would allow studies such as this one to correlate structural findings and persistence of symptoms that may be more subtle that findings detected by clinical evaluation.

One outstanding issue which this study does not resolve is the element of laterality in SCP abnormalities in children with CMS. Reports in healthy children^9^ and adults^44^ indicate no differences in FA or other diffusion MRI metrics between left and right DRTC tracts. However, many of the aforementioned studies have drawn different conclusions as to the required laterality of SCP damage in CMS; be it left^15^ – which our results are in agreement with – right^19^, or bilateral^12,14,36,37,45^. These differences may be due in part to the small sample sizes seen in many of these reports, and collaborative multi-centre approaches to tackle this problem may provide more definitive answers.

## Conclusions

This study adds significantly to existing evidence implicating damage to the superior cerebellar peduncle in cerebellar mutism syndrome after resection of medulloblastoma. In a richly-sampled group of children with medulloblastoma and age- and sex-matched healthy controls, along-tract profilometry was used to depict changes in FA in the cerebellar peduncles at a heretofore unprecedented level of spatiotemporal detail. Pre-operative FA appears to be insufficiently sensitive as a biomarker of CMS risk. Children with post-operative CMS showed focal changes in the distal regions of the left SCP, which were seen up to a year post-operatively. By pinpointing a specific region of the cerebellar white matter implicated in CMS, these findings have direct clinical implications for neurosurgeons performing resection of midline paediatric posterior fossa tumours.

## Data Availability

Anonymised data reported within this paper are available upon reasonable request to the senior author

## Acknowledgements

The authors would like to thank the children and their families who took part in this study, and to all staff in the Radiology Department for facilitating research scans in patients at Lucile Packard Children’s Hospital. The authors would also like to thank Virginia Marchman and Jonathan Clayden for advice on statistical analysis and data visualisation.

## Funding

SMT is funded by Great Ormond Street Hospital Children’s Charity (award no 174385), the UCL Bogue Fellowship, and is an Honorary Research Fellow of the Royal College of Surgeons of England. All research at Great Ormond Street Hospital NHS Foundation Trust and UCL Great Ormond Street Institute of Child Health is made possible by the NIHR Great Ormond Street Hospital Biomedical Research Centre. Stanford Children’s Health-Lucile Packard Children’s Hospital Center for Brain and Behavior Awards in Pediatric Neurosciences provided funding support for KWY. The National Institutes of Health provided funding for KET and HMF (HD084749 HD06915).

## Competing interests

The authors do not have any competing interests to disclose.

## Supplementary Material

**Supplementary Figure 1.**
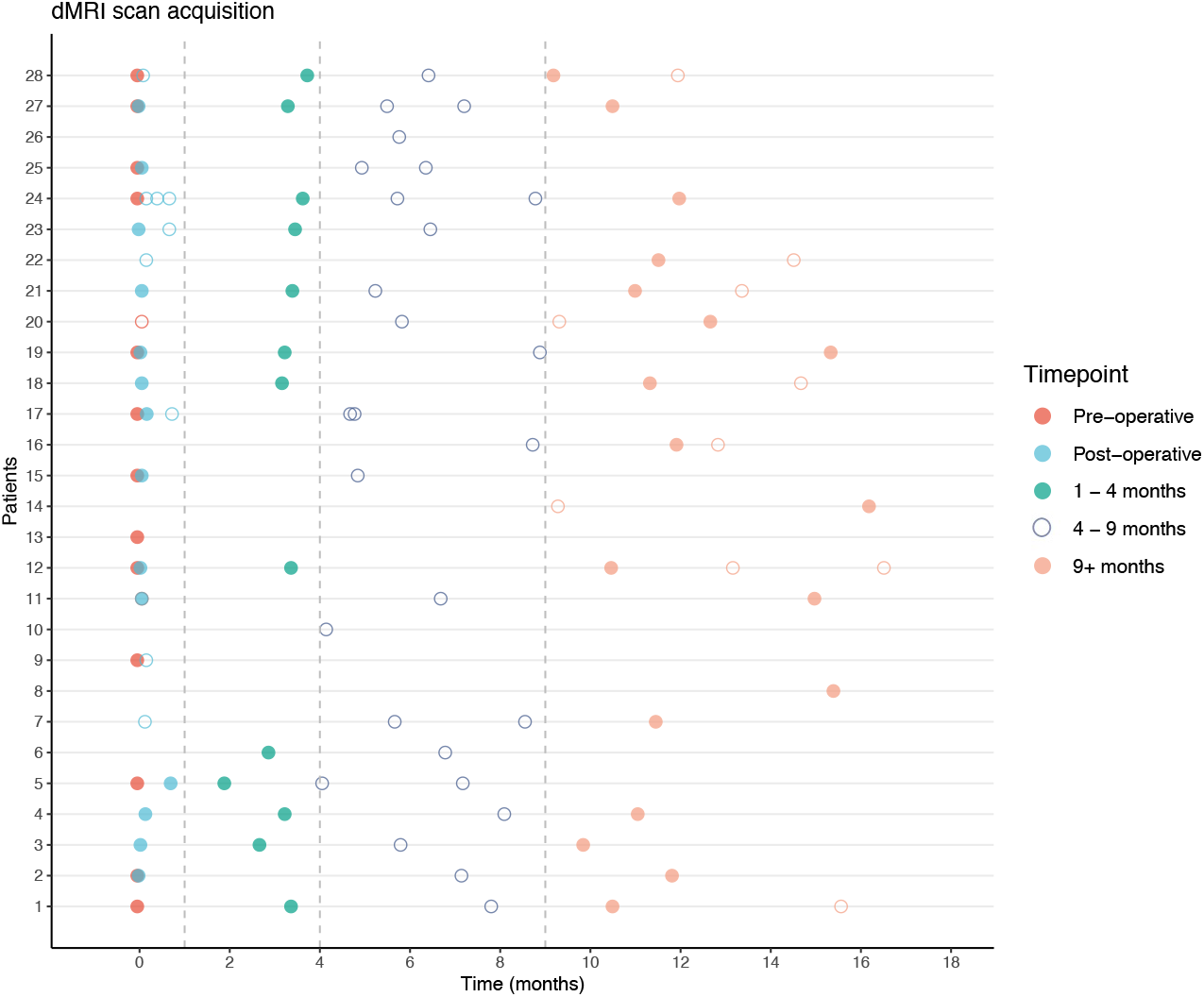
Diffusion MRI scan acquisition for eligible subjects (n=28). Scans were grouped into four timepoints: pre-operative, post-operative, early follow-up (1-4 months) and late follow-up (>9 months). Scans acquired between 4 and 9 months post-operatively (dark blue circles) were not included in the analysis. Filled circles indicate scan inclusion in analysis.

**Supplementary Table 1.**
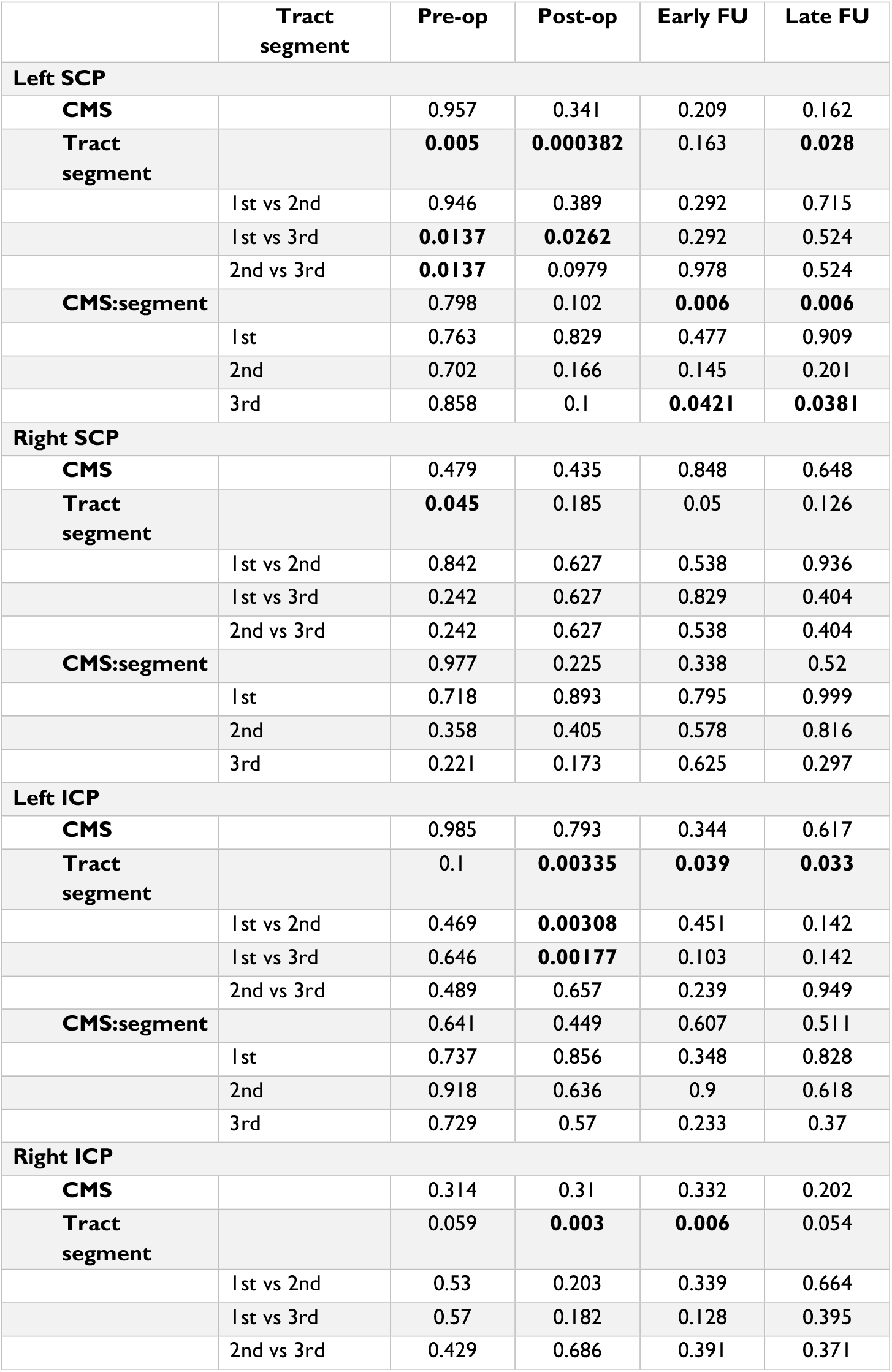
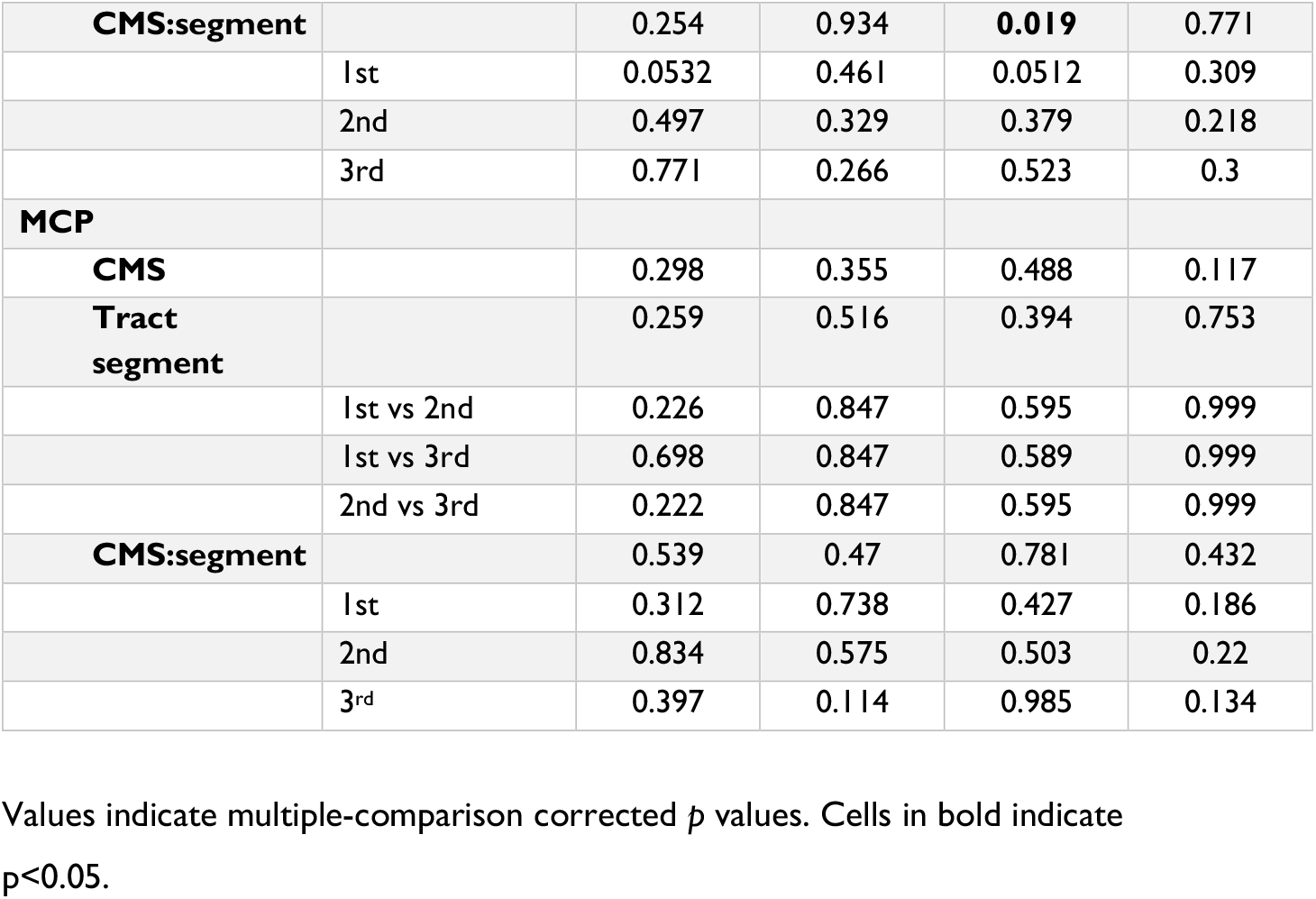
Fractional Anisotropy Z-score mixed ANOVA results.

|  | Tract segment | Pre-op | Post-op | Early FU | Late FU |
| --- | --- | --- | --- | --- | --- |
| **Left SCP** |  |  |  |  |  |
| **CMS** |  | 0.957 | 0.341 | 0.209 | 0.162 |
| **Tract segment** |  | **0.005** | **0.000382** | 0.163 | **0.028** |
|  | 1st vs 2nd | 0.946 | 0.389 | 0.292 | 0.715 |
|  | 1st vs 3rd | **0.0137** | **0.0262** | 0.292 | 0.524 |
|  | 2nd vs 3rd | **0.0137** | 0.0979 | 0.978 | 0.524 |
| **CMS:segment** |  | 0.798 | 0.102 | **0.006** | **0.006** |
|  | 1st | 0.763 | 0.829 | 0.477 | 0.909 |
|  | 2nd | 0.702 | 0.166 | 0.145 | 0.201 |
|  | 3rd | 0.858 | 0.1 | **0.0421** | **0.0381** |
| **Right SCP** |  |  |  |  |  |
| **CMS** |  | 0.479 | 0.435 | 0.848 | 0.648 |
| **Tract segment** |  | **0.045** | 0.185 | 0.05 | 0.126 |
|  | 1st vs 2nd | 0.842 | 0.627 | 0.538 | 0.936 |
|  | 1st vs 3rd | 0.242 | 0.627 | 0.829 | 0.404 |
|  | 2nd vs 3rd | 0.242 | 0.627 | 0.538 | 0.404 |
| **CMS:segment** |  | 0.977 | 0.225 | 0.338 | 0.52 |
|  | 1st | 0.718 | 0.893 | 0.795 | 0.999 |
|  | 2nd | 0.358 | 0.405 | 0.578 | 0.816 |
|  | 3rd | 0.221 | 0.173 | 0.625 | 0.297 |
| **Left ICP** |  |  |  |  |  |
| **CMS** |  | 0.985 | 0.793 | 0.344 | 0.617 |
| **Tract segment** |  | 0.1 | **0.00335** | **0.039** | **0.033** |
|  | 1st vs 2nd | 0.469 | **0.00308** | 0.451 | 0.142 |
|  | 1st vs 3rd | 0.646 | **0.00177** | 0.103 | 0.142 |
|  | 2nd vs 3rd | 0.489 | 0.657 | 0.239 | 0.949 |
| **CMS:segment** |  | 0.641 | 0.449 | 0.607 | 0.511 |
|  | 1st | 0.737 | 0.856 | 0.348 | 0.828 |
|  | 2nd | 0.918 | 0.636 | 0.9 | 0.618 |
|  | 3rd | 0.729 | 0.57 | 0.233 | 0.37 |
| **Right ICP** |  |  |  |  |  |
| **CMS** |  | 0.314 | 0.31 | 0.332 | 0.202 |
| **Tract segment** |  | 0.059 | **0.003** | **0.006** | 0.054 |
|  | 1st vs 2nd | 0.53 | 0.203 | 0.339 | 0.664 |
|  | 1st vs 3rd | 0.57 | 0.182 | 0.128 | 0.395 |
|  | 2nd vs 3rd | 0.429 | 0.686 | 0.391 | 0.371 |
| **CMS:segment** |  | 0.254 | 0.934 | **0.019** | 0.771 |
|  | 1st | 0.0532 | 0.461 | 0.0512 | 0.309 |
|  | 2nd | 0.497 | 0.329 | 0.379 | 0.218 |
|  | 3rd | 0.771 | 0.266 | 0.523 | 0.3 |
| **MCP** |  |  |  |  |  |
| **CMS** |  | 0.298 | 0.355 | 0.488 | 0.117 |
| **Tract segment** |  | 0.259 | 0.516 | 0.394 | 0.753 |
|  | 1st vs 2nd | 0.226 | 0.847 | 0.595 | 0.999 |
|  | 1st vs 3rd | 0.698 | 0.847 | 0.589 | 0.999 |
|  | 2nd vs 3rd | 0.222 | 0.847 | 0.595 | 0.999 |
| **CMS:segment** |  | 0.539 | 0.47 | 0.781 | 0.432 |
|  | 1st | 0.312 | 0.738 | 0.427 | 0.186 |
|  | 2nd | 0.834 | 0.575 | 0.503 | 0.22 |
|  | 3rd | 0.397 | 0.114 | 0.985 | 0.134 |

Values indicate multiple-comparison corrected $p$ values. Cells in bold indicate $p < 0.05$ .

## Notes

### Competing Interest Statement

The authors have declared no competing interest.

### Funding Statement

SMT is funded by Great Ormond Street Hospital Childrens Charity (award no 174385), the UCL Bogue Fellowship, and is an Honorary Research Fellow of the Royal College of Surgeons of England. All research at Great Ormond Street Hospital NHS Foundation Trust and UCL Great Ormond Street Institute of Child Health is made possible by the NIHR Great Ormond Street Hospital Biomedical Research Centre.

### Author Declarations

Stanford University School of Medicine / Lucile Packard Childrens Hospital Institutional Review Board #38471 #36206

